# CARDIOGENIC SHOCK MORTALITY ACCORDING TO AETIOLOGY IN A MEDITERRANEAN COHORT: RESULTS FROM THE SHOCK-CAT STUDY

**DOI:** 10.1101/2024.06.30.24309558

**Authors:** Cosme García-García, Teresa López-Sobrino, Esther Sanz-Girgas, Maria Ruiz Cueto, Jaime Aboal, Pablo Pastor, Irene Buera, Alessandro Sionis, Rut- Andrea, Judit Rodríguez-López, JC Sánchez-Salado, Carlos Tomas Querol, Jordi Bañeras, Albert Ariza Sole, Josep Lupón, Antoni Bayés-Genís, Ferran Rueda, Grup de Treball de Cures Agudes Cardiològiques Societat Catalana de Cardiologia

**Author notes:** Corresponding Author: Cosme Garcia-Garcia, MD, PhD. Heart Institute. Hospital Universitari Germans Trias i Pujol. Associate Professor Autonomous University of Barcelona Carretera de Canyet s/n 08916. Badalona (Barcelona). Twitter: @CosmeGarciacg7. Medical and translational research PhD Program. University of Barcelona. This author takes responsibility for all aspects of the reliability and freedom from bias of the data presented and their discussed interpretation.

## Abstract

**Background and objectives:** Mortality in cardiogenic shock (CS) remains elevated, with the potential for CS causes to impact prognosis and risk stratification. The aim was to investigate in-hospital prognosis and mortality in CS patients according to aetiology. We also assessed the prognostic accuracy of CardShock and IABP-SHOCK II scores.

**Methods:** Shock-CAT study was a multicentre, prospective, observational study conducted from December 2018-November 2019 in eight University hospitals in Catalonia, including non-selected consecutive CS patients. Data on clinical presentation, management, including mechanical circulatory support (MCS) were analysed comparing acute myocardial infarction (AMI) related CS and non-AMI-CS. The accuracy of CardShock and IABP-SHOCK II scores to assess 90-days mortality risk were also compared.

**Results:** A total of 382 CS patients were included, age 65.3 (SD 13.9) years, 75.1% men. Patients were classified as AMI-CS (n=232, 60.7%) and non-AMI-CS (n=150, 39.3%). In the AMI-CS group, 77.6% were STEMI. Main aetiologies for non-AMI-CS were heart failure (36.2%), arrhythmias (22.1%) and valve disease (8.0%). AMI-CS patients required more MCS than non-AMI-CS (43.1% vs 16.7%, p<0.001). In-hospital mortality was higher in AMI-CS (37.1 vs 26.7%, p=0.035), with a two-fold increased risk after multivariate adjustment (OR 2.24, p=0.019). The IABP-SHOCK II had superior discrimination for predicting 90-days mortality when compared with CardShock in AMI-CS patients (AUC 0.74 vs 0.66, p=0.047) although both scores performed similarly in non-AMI-CS (AUC 0.64 vs 0.62, p=0.693).

**Conclusions:** In our cohort, AMI-CS mortality was increased by two-fold when compared to non- AMI-CS. IABP-SHOCK II score provides better 90-days mortality risk prediction than CardShock score in AMI-CS, but both scores performed similar in non-AMI-CS patients.

## ABBREVIATIONS

**ABBREVIATIONS:**
|  |  |
| --- | --- |
| CS | Cardiogenic Shock |
| MCS | Mechanical circulatory support |
| AMI | Acute myocardial infarction |
| STEMI | ST-Elevation acute myocardial infarction |
| ICCU | Intensive Cardiac Care Unit |
| ECMO | Extracorporeal membrane oxygenation |

## ABREVIATURAS

**ABREVIATURAS:**
|  |  |
| --- | --- |
| SC | Shock Cardiogénico |
| SCM | Soporte Circulatorio Mecánico |
| IAM | Infarto agudo de miocardio |
| IAMEST | Infarto agudo de miocardio con elevación del ST |
| UCAC | Unidad de cuidados agudos cardiológicos |
| ECMO | Membrana de oxigenación extracorpórea |

## BACKGROUND

Cardiogenic shock (CS) is a critical condition consisting in severe systemic hypoperfusion due to primary cardiac dysfunction (1,2). Despite the advances in acute cardiac care, from reperfusion to mechanical circulatory support (MCS), mortality from CS remains very high (3,4).

Acute myocardial infarction (AMI) stands as the primary contributor to cardiogenic shock (CS) (5–7). Despite reports from certain studies in the United States indicating a rise in CS incidence (3,8), a recent Spanish registry spanning three decades within a Mediterranean cohort revealed no alterations in the prevalence of CS stemming from ST-elevation AMI (STEMI) (5). Nevertheless, the improved prognosis of AMI, highlighted by a notable reduction in acute-phase mortality (9), coupled with an increase in the prevalence of chronic heart failure patients (10), might have reshaped the clinical profile and underlying causes of CS patients. A similar trend was observed in an American CS registry, noting a 30% reduction in CS cases complicated by AMI between 2005 and 2014 (11). However, despite the conjectured alterations in aetiology, there exists limited information regarding the potential impact of aetiology on both short-term and medium-term mortality rates in CS patients. Furthermore, variations in aetiology could potentially lead to changes in treatment strategies, including the utilization of MCS and other invasive procedures. Presently, there is an absence of data from randomized trials that specifically examine the management of CS based on etiological factors.

Furthermore, there is an enduring interest in effectively stratifying the risk of CS patients, aiming to customize the most suitable treatments, including the timely implementation of MCS for those at a higher risk. In addition, it is worth noting that the underlying cause of the shock might have an impact on the predictive precision of the most valuable risk assessment scores accessible for these patients (12,13).

The primary objective of the Shock CAT registry was to evaluate the in-hospital prognosis and acute phase and mid-term mortality risk among CS patients, contingent upon the underlying cause of the shock. The study involved an analysis of the clinical characteristics of an unselective and real-world cohort of CS patients from the Mediterranean region. This analysis encompassed a comparison of the treatment strategies, including the utilization of MCS, as well as the examination of 90-day and 6- month mortality rates between patients experiencing AMI and those with non-AMI-related cardiogenic shock. The accuracy of the CardShock and IABP-SHOCK II scores (12,13) was also analysed to asses 90-days mortality risk in both groups.

## METHODS

### Study population

The Shock-CAT study is a regional prospective, observational multicentre study on CS. Patients were recruited from 1^st^ December 2018 and 30^th^ November 2019 from intensive cardiac care units (ICCU) of eight University hospitals in Catalonia (Spain). All these hospitals had an own ICCU, conducted by cardiologist with specific formation in the management of critical cardiac patients. Moreover, all eight hospitals had cath lab 24/7 availability and most of them (75%) had an own cardiac surgery service and extracorporeal membrane oxygenation (ECMO) availability in the same center. The study enrolled consecutive patients aged over 18 years with CS of different aetiologies. The CS could be present at hospital admission or developed during in-hospital stay. CS was defined following the classical and guidelines definitions of CS (2,14), as systolic blood pressure <90 mm Hg (after adequate fluid challenge) for 30 minutes or a need for vasopressor therapy to maintain systolic blood pressure >90 mm Hg, and signs of hypoperfusion (altered mental status/confusion, cold periphery, oliguria <0.5mL/kg/h for the previous 6 h, or blood lactate >2 mmol/L). Patients developing CS after cardiac or non-cardiac surgery were excluded. The aetiology of CS was determined by local investigators depending on the main diagnosis at admission. Patients were classified in two groups, depending on the CS was due to an AMI or other causes (non-AMI). CS management was performed according to local physician’s criteria, following the current guidelines and recommendations (15,16). Baseline demographics and clinical data were recorded during hospital admission in a database. Outcome events were adjudicated based on electronic clinical records and/or directly contacting patients or relatives by telephone.

The study was approved by the local Ethics Committee and conducted in accordance with the biomedical research law 14/2007 within the Helsinki Declaration framework. All patients provided written consent by signing the specific consent form of the study. If patients were unconscious and unable to consent, therefore informed consent was obtained from their relatives.

### Statistical analysis

Results are presented as number (n) and percentages (%). Categorical variables are expressed as frequencies and percentages and continuous variables as means and standard deviation or medians and interquartile range depending on the normal or not-normal variable distribution. Departures from normality were evaluated using normal QQ-plots. Statistical differences between groups were compared using the χ^2^ and Student’s t-test and Mann-Whitney U test. Multivariate analysis was performed with logistic regression models, with different covariates to identify variables associated with CS in-hospital mortality. A sensitivity analysis after propensity score matching was performed including ECMO capability hospital and all variables used in the multivariate analysis to assess in-hospital mortality. Assumptions of linearity of continuous variables (lineal regression) were tested. Probability values <0.05 from two-sided tests were considered to indicate statistical significance. All analyses were performed using the software IBM Statistics SPSS 24 (Chicago, Illinois, USA) and STATA 17.

## RESULTS

### Study population

A total of 382 patients were included in the Shock-CAT. Mean age was 65.3 (SD 14) years and 24.9% (95 patients) were women. AMI was the main cause of CS in 232 patients (60.7%) while 150 patients (39.3%) formed the non-AMI-CS group. The main aetiology in AMI-CS group was STEMI in 77.6%, whereas in non-AMI-CS, heart failure was the main diagnose in 36.2% of them, followed by malignant arrhythmias (22.1%), severe aortic stenosis (8%) and myocarditis (7.4%). All diagnoses at admission in both groups are shown in Figure 1. Out-of-hospital cardiac arrest occurred in 133 patients (34.8%) with a slightly higher proportion in AMI-CS patients (38.4% vs 30%).

**Figure 1:**
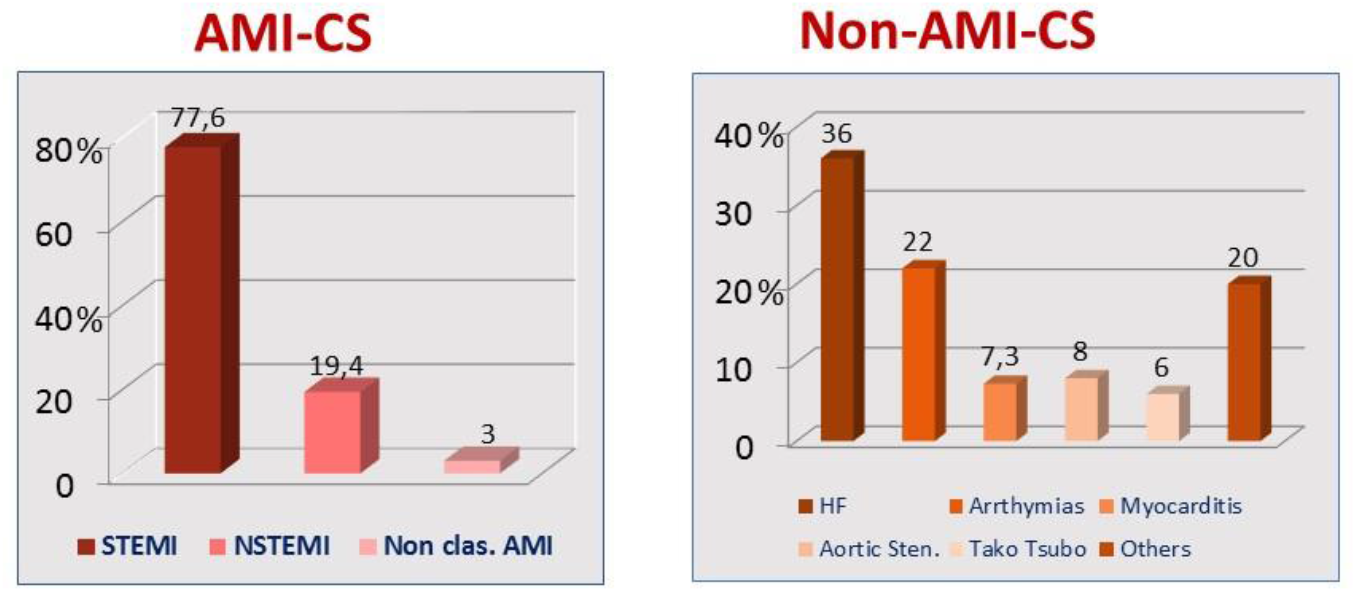
Etiologies of Cardiogenic shock in Shock-CAT registry in AMI-CS (left) and non-AMI-CS (right).

Baseline characteristics are described in Table 1. Briefly, non-AMI-CS patients tended to be younger than AMI-CS patients (63.9 vs 66.2 years) and were more frequently women (36% vs 17.7%). Cardiovascular risk factors were prevalent although there were not differences between both groups. In the non-AMI-CS group there was a higher proportion of patients with previous history of heart failure (40% vs 4.1%) and prior MI (24.8% vs 13.9%). Haemodynamic and biochemical parameters were similar between groups, both systolic and diastolic blood pressure, as well as heart rate, without differences in lactate peak or shock index (ratio between heart rate and systolic blood pressure). Mean left ventricle ejection fraction was 31.8%, slightly higher in AMI-CS patients (32.9% vs 30.1%, p=0.05). Non-AMI-CS patients had worse renal function.

**Table 1.**
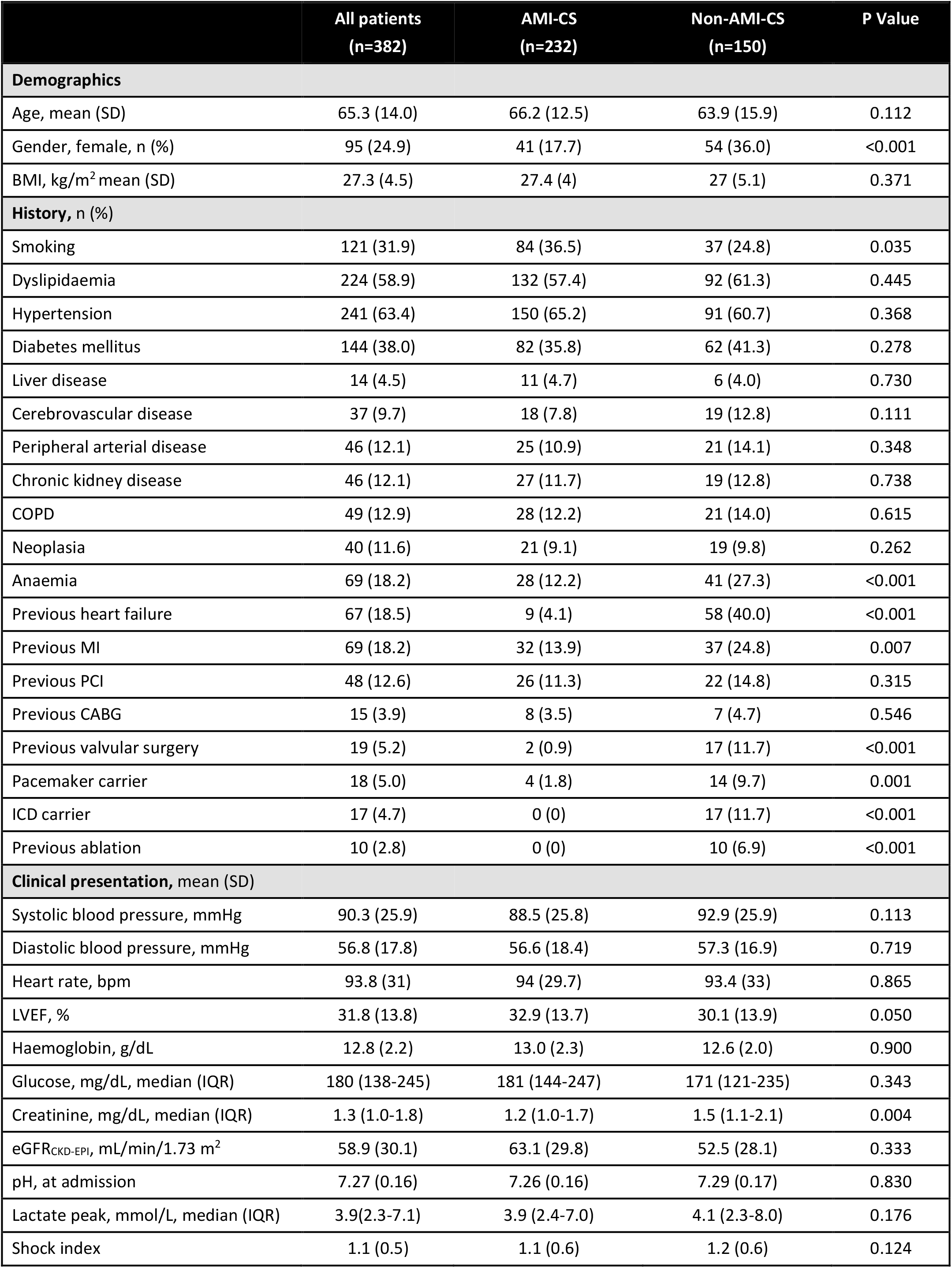
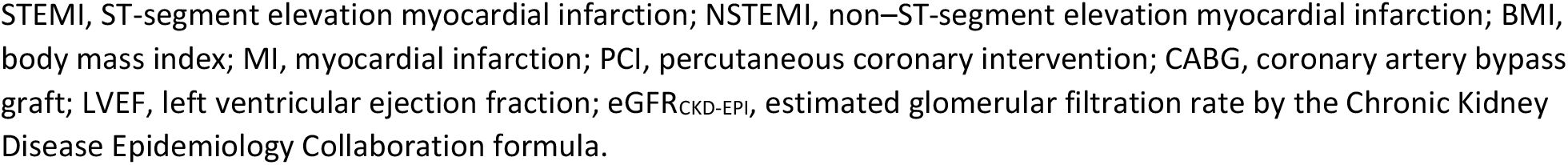
Baseline characteristics of the study population by presentation as AMI-CS or non-AMI-CS.

|  | All patients<br>(n=382) | AMI-CS<br>(n=232) | Non-AMI-CS<br>(n=150) | P Value |
| --- | --- | --- | --- | --- |
| **Demographics** |  |  |  |  |
| Age, mean (SD) | 65.3 (14.0) | 66.2 (12.5) | 63.9 (15.9) | 0.112 |
| Gender, female, n (%) | 95 (24.9) | 41 (17.7) | 54 (36.0) | <0.001 |
| BMI, kg/m<sup>2</sup> mean (SD) | 27.3 (4.5) | 27.4 (4) | 27 (5.1) | 0.371 |
| **History, n (%)** |  |  |  |  |
| Smoking | 121 (31.9) | 84 (36.5) | 37 (24.8) | 0.035 |
| Dyslipidaemia | 224 (58.9) | 132 (57.4) | 92 (61.3) | 0.445 |
| Hypertension | 241 (63.4) | 150 (65.2) | 91 (60.7) | 0.368 |
| Diabetes mellitus | 144 (38.0) | 82 (35.8) | 62 (41.3) | 0.278 |
| Liver disease | 14 (4.5) | 11 (4.7) | 6 (4.0) | 0.730 |
| Cerebrovascular disease | 37 (9.7) | 18 (7.8) | 19 (12.8) | 0.111 |
| Peripheral arterial disease | 46 (12.1) | 25 (10.9) | 21 (14.1) | 0.348 |
| Chronic kidney disease | 46 (12.1) | 27 (11.7) | 19 (12.8) | 0.738 |
| COPD | 49 (12.9) | 28 (12.2) | 21 (14.0) | 0.615 |
| Neoplasia | 40 (11.6) | 21 (9.1) | 19 (9.8) | 0.262 |
| Anaemia | 69 (18.2) | 28 (12.2) | 41 (27.3) | <0.001 |
| Previous heart failure | 67 (18.5) | 9 (4.1) | 58 (40.0) | <0.001 |
| Previous MI | 69 (18.2) | 32 (13.9) | 37 (24.8) | 0.007 |
| Previous PCI | 48 (12.6) | 26 (11.3) | 22 (14.8) | 0.315 |
| Previous CABG | 15 (3.9) | 8 (3.5) | 7 (4.7) | 0.546 |
| Previous valvular surgery | 19 (5.2) | 2 (0.9) | 17 (11.7) | <0.001 |
| Pacemaker carrier | 18 (5.0) | 4 (1.8) | 14 (9.7) | 0.001 |
| ICD carrier | 17 (4.7) | 0 (0) | 17 (11.7) | <0.001 |
| Previous ablation | 10 (2.8) | 0 (0) | 10 (6.9) | <0.001 |
| **Clinical presentation, mean (SD)** |  |  |  |  |
| Systolic blood pressure, mmHg | 90.3 (25.9) | 88.5 (25.8) | 92.9 (25.9) | 0.113 |
| Diastolic blood pressure, mmHg | 56.8 (17.8) | 56.6 (18.4) | 57.3 (16.9) | 0.719 |
| Heart rate, bpm | 93.8 (31) | 94 (29.7) | 93.4 (33) | 0.865 |
| LVEF, % | 31.8 (13.8) | 32.9 (13.7) | 30.1 (13.9) | 0.050 |
| Haemoglobin, g/dL | 12.8 (2.2) | 13.0 (2.3) | 12.6 (2.0) | 0.900 |
| Glucose, mg/dL, median (IQR) | 180 (138-245) | 181 (144-247) | 171 (121-235) | 0.343 |
| Creatinine, mg/dL, median (IQR) | 1.3 (1.0-1.8) | 1.2 (1.0-1.7) | 1.5 (1.1-2.1) | 0.004 |
| eGFR<sub>CKD-EPI</sub>, mL/min/1.73 m<sup>2</sup> | 58.9 (30.1) | 63.1 (29.8) | 52.5 (28.1) | 0.333 |
| pH, at admission | 7.27 (0.16) | 7.26 (0.16) | 7.29 (0.17) | 0.830 |
| Lactate peak, mmol/L, median (IQR) | 3.9(2.3-7.1) | 3.9 (2.4-7.0) | 4.1 (2.3-8.0) | 0.176 |
| Shock index | 1.1 (0.5) | 1.1 (0.6) | 1.2 (0.6) | 0.124 |

STEMI, ST-segment elevation myocardial infarction; NSTEMI, non-ST-segment elevation myocardial infarction; BMI, body mass index; MI, myocardial infarction; PCI, percutaneous coronary intervention; CABG, coronary artery bypass graft; LVEF, left ventricular ejection fraction; $\text{eGFR}_{\text{CKD-EPI}}$ , estimated glomerular filtration rate by the Chronic Kidney Disease Epidemiology Collaboration formula.

Management and procedures during hospital admission are detailed in Table 2. Overall, 36.1% of patients received MCS, with a higher proportion in AMI-CS patients (43.5% vs 24.5%, p<0.001), that was maintained for all types of percutaneous short term MCS such as intraaortic balloon pump (39.3% vs 16.4%) and Impella CP® (15.7% vs 6.5%). Extracorporeal membrane oxygenation (ECMO) was performed in 9.8% (36 patients) with a trend to higher proportion in AMI-CS patients (11.7% vs 6.5%, p=0.096). Only 16 patients (4.3%) underwent heart transplant without differences between groups. Almost all patients (97.4%) with AMI-CS aetiology underwent coronary angiogram, and 83.2% of these underwent percutaneous coronary intervention (PCI). Reperfusion therapy was successfully achieved in 84.3% of STEMI patients, all with primary PCI. In non-AMI-CS patients, coronary angiography was done only in 68.3% of patients and nearby two thirds of them had coronary arteries without obstructive lesions. Invasive monitoring with pulmonary artery catheter was performed in 27.7% of patients, without differences between groups.

**Table 2.** ICCU procedures and medical therapies.

|  | All patients<br>(n=382) | AMI-CS<br>(n=232) | Non-AMI-CS<br>(n=150) | P Value |
| --- | --- | --- | --- | --- |
| **Catheterization lab data, n (%)** |  |  |  |  |
| Coronary angiography | 323 (86.1) | 224 (97.4) | 99 (68.3) | <0.001 |
| Percutaneous Coronary intervention | 210 (55) | 193 (83.2) | 17 (11.3) | <0.001 |
| Main epicardial coronary arteries ≥70% stenosis |  |  |  | <0.001 |
| 0 | 67(17.8) | 4 (1.8) | 63 (63.6) |  |
| 1 | 101 (26.9) | 83 (37.1) | 18 (18.2) |  |
| 2 | 70 (18.6) | 59 (26.3) | 11 (11.1) |  |
| 3 | 99 (26.3) | 76 (33.9) | 23 (23.2) |  |
| Left main ≥ 50% stenosis | 52 (13.6) | 45 (20.1) | 7 (7.1) |  |
| Reperfusion therapy (in STEMI patients) | ---- | 150 (84.3) | ---- | ---- |
| **ICCU procedures and treatments, n (%)** |  |  |  |  |
| Mechanical ventilation |  |  |  |  |
| Invasive | 241 (63.9) | 151 (65.7) | 90 (61.2) | 0.383 |
| Non-invasive | 112 (30.1) | 61 (27.1) | 51 (34.7) | 0.119 |
| Days with mechanical ventilation | 9.5 (11.5) | 9.6 (12.4) | 9. (12.2) | 0.844 |
| Renal replacement therapy | 51 (13.6) | 29 (12.6) | 22 (15.2) | 0.481 |
| Hypothermia | 74 (19.6) | 47 (20.4) | 27 (18.4) | 0.622 |
| Temporary pacemaker | 69 (18.4) | 43 (18.8) | 26 (17.7) | 0.790 |
| Pulmonary artery catheter | 103 (27.7) | 61 (26.9) | 42 (29) | 0.660 |
| Inotropes' | 361 (96.3) | 224 (98.2) | 137 (93.2) | 0.012 |
| Days with inotrope's | 6.2 (9.1) | 6.3 (10.3) | 6.1 (6.7) | 0.875 |
| Ventricular Assist Devices | 136 (36.1) | 100 (43.5) | 36 (24.5) | <0.001 |
| IABP | 114 (30.4) | 90 (39.3) | 24 (16.4) | <0.001 |
| Impella CP | 45 (12.2) | 36 (15.7) | 9 (6.5) | 0.008 |
| ECMO | 36 (9.8) | 27 (11.7) | 9 (6.5) | 0.096 |
| Centrimag Biventricular | 16 (4.3) | 7 (3.1) | 9 (6.2) | 0.147 |
| Centrimag univentricular | 4 (1.1) | 3 (1.3) | 1 (0.7) | 0.566 |
| Cardiac transplant | 16 (4.3) | 7 (3.1) | 9 (6.2) | 0.262 |
| CABG | 43 (11.7) | 22 (9.9) | 21 (14.6) | 0.328 |
| Computed Tomography | 152 (40.6) | 81 (35.5) | 71 (48.6) | 0.012 |
| Red blood cells transfusion | 110 (30.1) | 64 (28.6) | 46 (32.4) | 0.437 |
| Magnetic Resonance | 64 (17.0) | 28 (12.2) | 36 (24.5) | 0.002 |
| **Admission to ECMO Capability Hospital** | **285 (74.6)** | **167 (72)** | **118 (78.7)** | **0.143** |

Relative to medical therapies, inotropes or vasopressors were used in the majority of patients (96.3%), with more frequent use of dobutamine in AMI patients (86.4% vs 68%, p<0.001) and levosimendan in non-AMI (12.9% vs 5.7%, p=0.015); norepinephrine was the main vasopressor, administered to 77.6% of all patients, without differences between groups. Aspirin, other antiplatelet therapies and statins were more often used in AMI than non-AMI, as expected. Medical treatments are detailed in supplementary Table 1. All CS patients suffered many in-hospital complications: mainly infection in more than half of patients or major bleeding in 11.2%, which did not differ between groups, a higher proportion of acute kidney injury in non-AMI-CS patients (64.8% vs. 49.3%), and ventricular fibrillation and mechanical complications were more frequent in AMI-CS group than in non-AMI-CS. In the AMI group, mechanical complications occurred in 9% of patients (6 patients with free wall rupture, 8 ventricular septal rupture and 7 papillary muscle rupture). In-hospital complications are shown in Table 3.

**Table 3.** In-hospital complications, acute-phase mortality and mid-term outcomes. Risk scores by aetiology.

|  | All patients<br>(n=382) | AMI-CS<br>(n=232) | Non-AMI CS<br>(n=150) | P Value |
| --- | --- | --- | --- | --- |
| **MI complications, n (%)** |  |  |  |  |
| OHCA, at admission | 133 (34.8) | 88 (38.4) | 45 (30) | 0.091 |
| -ROSC delay (minutes) | 27.7 (20.1) | 28.1 (20.4) | 27 (19.7) | 0.764 |
| Ventricular fibrillation | 56 (15.2) | 44 (19.4) | 12 (8.5) | 0.004 |
| Sustained monomorphic ventricular tachycardia | 55 (14.9) | 35 (15.4) | 20 (14.1) | 0.726 |
| 3<sup>rd</sup> degree atrioventricular block | 45 (12.2) | 32 (14.1) | 13 (9.2) | 0.158 |
| Right Ventricle AMI | ---- | 18 (7.9) | ----- | ----- |
| Atrial fibrillation | 90 (22.4) | 53 (23.3) | 37 (26.1) | 0.556 |
| Acute conduction disturbance | 6 (1.6) | 6 (2.6) | 0 (0) | 0.051 |
| **Mechanical complications** |  |  |  |  |
| Free wall rupture | 7 (1.9) | 6 (2.6) | 1 (0.7) | 0.184 |
| Ventricular septal rupture | 9 (2.4) | 8 (3.5) | 1 (0.7) | 0.088 |
| Papillary muscle rupture | 8 (2.2) | 7 (3.1) | 1 (0.7) | 0.127 |
| Stroke | 18 (4.9) | 16 (7.2) | 2 (1.4) | 0.145 |
| Major bleeding | 41 (11.2) | 27 (12.1) | 14 (9.7) | 0.795 |
| Infection | 194 (52.4) | 118 (52.7) | 76 (52.1) | 0.906 |
| Acute kidney injury | 204 (55.4) | 110 (49.3) | 94 (64.8) | 0.013 |
| **Discharge, mean (SD)** |  |  |  |  |
| Length of ACCU admission, days | 14.0 (16.5) | 13.1 (5.7) | 15.4 (17.7) | 0.187 |
| Length of hospital admission, days | 22.0 (24.7) | 20.9 (25.6) | 23.7 (23.8) | 0.289 |
| **Prognosis, n (%)** |  |  |  |  |
| In-hospital mortality | 126 (33) | 86 (37.1) | 40 (26.7) | 0.035 |
| 90-days mortality | 141 (37.3) | 94 (40.9) | 47 (31.8) | 0.074 |
| 6-month mortality | 157 (41.5) | 104 (45.2) | 53 (35.8) | 0.176 |
| 6-month readmission (Cardiovascular) | 22 (8.7) | 14 (9.7) | 8 (7.3) | 0.155 |
| **Risk scores, mean (SD)** |  |  |  |  |
| CardShock | 4.3 (1.74) | 4.5 (1.71) | 4.0 (1.73) | 0.006 |
| IABP-SHOCK II | 2.2 (1.61) | 2.4 (1.65) | 1.9 (1.49) | 0.005 |
| INTERMACS stage | 2.3 (0.9) | 2.2 (0.9) | 2.4 (0.8) | 0.017 |
| GRACE Score | --- | 213 (53) | ---- |  |
| SCAI Classification, n (%) |  |  |  | 0.249 |
| B | 54 (14.2) | 26 (11.2) | 28 (18.7) |  |
| C | 189 (49.5) | 118 (50.9) | 71 (47.3) |  |
| D | 81 (21.2) | 50 (21.6) | 31 (20.7) |  |
| E | 58 (15.2) | 38 (16.4) | 20 (13.3) |  |

Shock-CAT registry was designed and performed before SCAI classification was published (17) although authors retrospectively collected SCAI classification data in our database. There were no differences in SCAI classification between AMI-CS and non-AMI CS (Table 3). SCAI D or E was observed in 36.4% of all CS patients (37.9% AMI-CS and 34% in non-AMI-CS, p= 0.435).

### Mortality rates and risk prediction

There were 126 in-hospital deaths, 33% of all CS patients. In-hospital mortality was higher in AMI-CS patients compared to non-AMI-CS group (37.1% vs 26.7%, p=0.035). The overall 90-days mortality was 37.3%, and was also trend higher in AMI-CS patients compared to those with non- AMI-CS (40.9% vs 31.8%, p=0.074). This difference in all-cause mortality remains among the first 6-months (Table 3), with non-significant trend to higher mortality in AMI patients (45.2% vs 35.8%). The 6-months follow up was completed in up to 98.9% of survivors. During this period an 8.7% of them were readmitted due to cardiovascular cause, without differences between groups.

Univariate analysis for in-hospital mortality showed that AMI-CS patients had higher risk than non-AMI-CS (OR 1.62; 95% CI 1.03 to 2.54, p=0.035). Moreover, multivariate Logistic Regression analyses confirmed that AMI-CS patients remained with a higher in-hospital mortality than non-AMI-CS after adjustment by age, gender, previous MI, previous heart failure, anemia, renal function, inotrope’s use, LVEF, MCS, SCAI grade D/E and ECMO capability hospital (OR 2.24, 95% CI 1.14 to 4.39, p=0.019) (Table 4). Moreover, a sensitivity analysis was performed in 208 patients after propensity score matching which showed that in-hospital mortality remains higher in AMI-CS patients than non-AMI-CS (40.4% vs 26.9%, p=0.04). The propensity score matching is shown in supplementary Table 2.

**Table 4.** Multivariate Logistic Regression analyses for in-hospital mortality.

|  | Univariate |  |  | Multivariate |  |  |
| --- | --- | --- | --- | --- | --- | --- |
|  | Odds Ratio | CI 95% | P Value | Odds Ratio | CI 95% | P Value |
| **Age** | **1.03** | **1.01-1.04** | **0.001** | **1.02** | **1.01-1.05** | **0.030** |
| Sex (women) | 1.11 | 0.68-1.81 | 0.675 | 1.18 | 0.61-2.29 | 0.626 |
| Previous MI | 1.50 | 0.88-2.56 | 0.140 | 1.30 | 0.62-2.69 | 0.487 |
| Previous Heart Failure | 0.91 | 0.52-1.61 | 0.753 | 1.02 | 0.43-2.39 | 0.971 |
| Anemia | 1.61 | 0.95-2.76 | 0.079 | 0.93 | 0.43-2.01 | 0.852 |
| **Creatinine at admission** | **2.64** | **1.90-3.68** | **<0.001** | **2.39** | **1.60-3.56** | **<0.001** |
| Left Ventricular Ejection Fraction | 0.99 | 0.97-1.01 | 0.219 | 0.99 | 0.97-1.01 | 0.530 |
| Inotrope's | 3.02 | 0.67-13.73 | 0.152 | 4.58 | 0.81-25.92 | 0.085 |
| MCS | 1.19 | 0.77-1.86 | 0.430 | 0.54 | 0.29-1.02 | 0.059 |
| **AMI-aetiology** | **1.62** | **1.03-2.54** | **0.035** | **2.24** | **1.14-4.39** | **0.019** |
| **SCAI D/E** | **6.89** | **4.30-11.06** | **<0.001** | **8.66** | **4.56-16.45** | **<0.001** |
| ECMO Capability Hospital | 1.21 | 0.73-1.99 | 0.454 | 0.71 | 0.37-1.38 | 0.315 |

AMI: Acute Myocardial Infarction; MI: Myocardial infarction; MCS: mechanical circulatory support

The CardShock and IABP-SHOCK II risk scores were applied to predict 90-days mortality in our cohort. Both scores showed a higher risk of death for AMI-CS patients than non-AMI-CS patients (Table 3). Moreover, AMI-CS group had also lower INTERMACS stage than non-AMI patients did. The prognostic accuracy of CardShock and IABP-SHOCK II scores was analysed, Receiver-operating characteristic curves demonstrated that IABP shock score had higher discrimination for predicting 90-days mortality when compared to Cardshock score in the entire cohort (area under the curve -AUC- 0.72 vs 0.66, p=0.042), Figure 2. Depending on the aetiology of CS, we found that IAPB Score had better accuracy for predicting 90-days mortality than CardShock in AMI patients (AUC 0.74 vs 0.66) respectively, p=0.047, although both scores were similar in non- AMI (AUC 0.64 vs 0.62, p=0.693), Figure 2.

**Figure 2.**
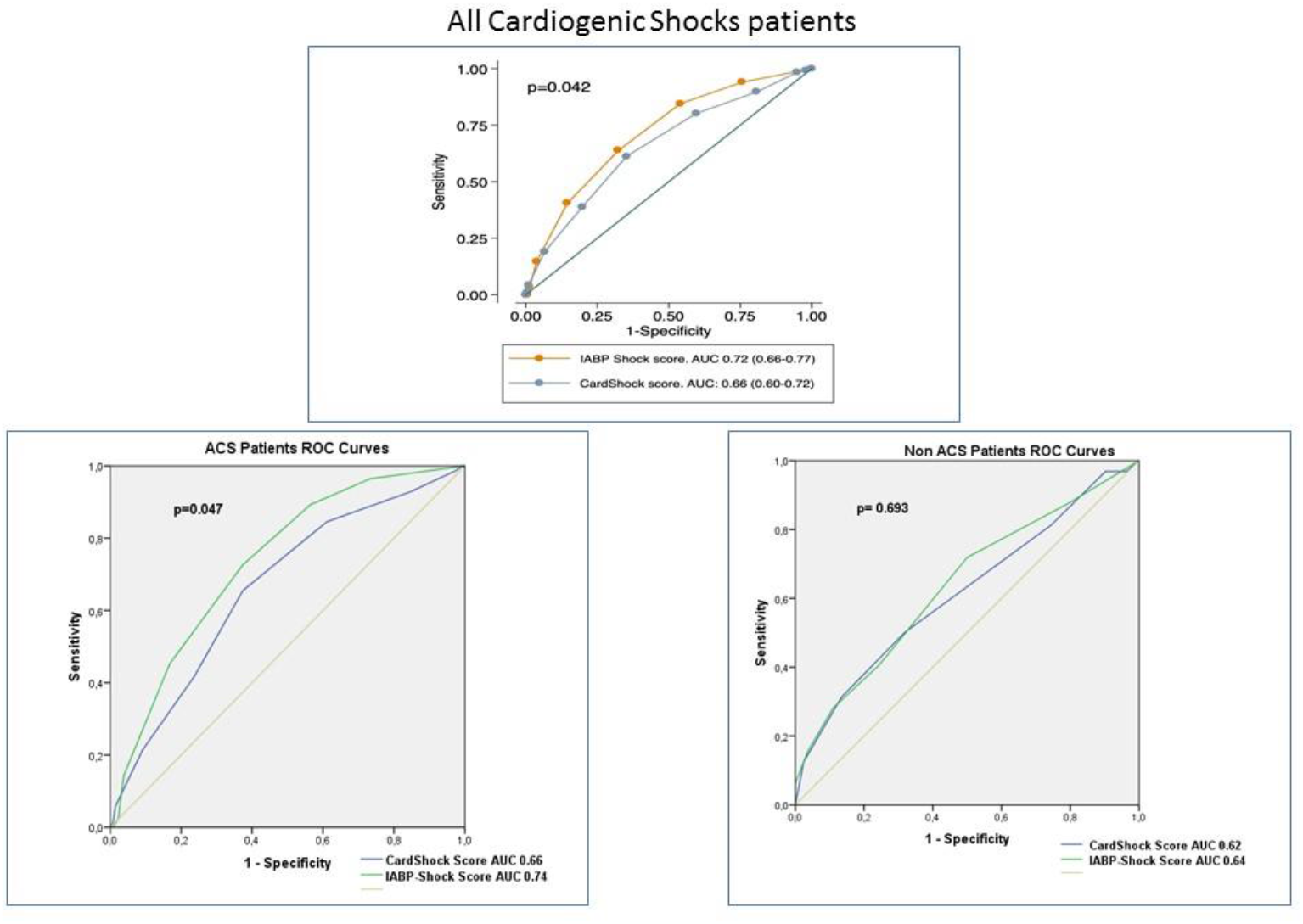
Cardshock and IABP Score accuracy for 90-days mortality in Shock-CAT registry. Top figure, all CS patients. Down figures: Depending on CS aetiology.

## DISCUSSION

The present study provides real-life data on acute-phase and mid-term prognosis in a non-selected Mediterranean cohort of CS patients depending on the aetiology of the shock. AMI remains the main cause of CS in above 60% of patients (being STEMI more than 75% of them). One third of CS patients died during hospital stay, with a higher rate of death in AMI-CS. After multivariate adjustment, the odds ratio for of in-hospital mortality was two-fold higher in AMI-CS than in non- AMI-CS. All-cause mortality remains with higher trend in AMI than non-AMI at 90-days and 6- months. IABP-SHOCK II score provided a better accuracy for predicting 90-days mortality in AMI-CS with respect to the CardShock, with both scores performing similarly in non-AMI-CS (Figure 3, Central Figure).

**Figure 3.**
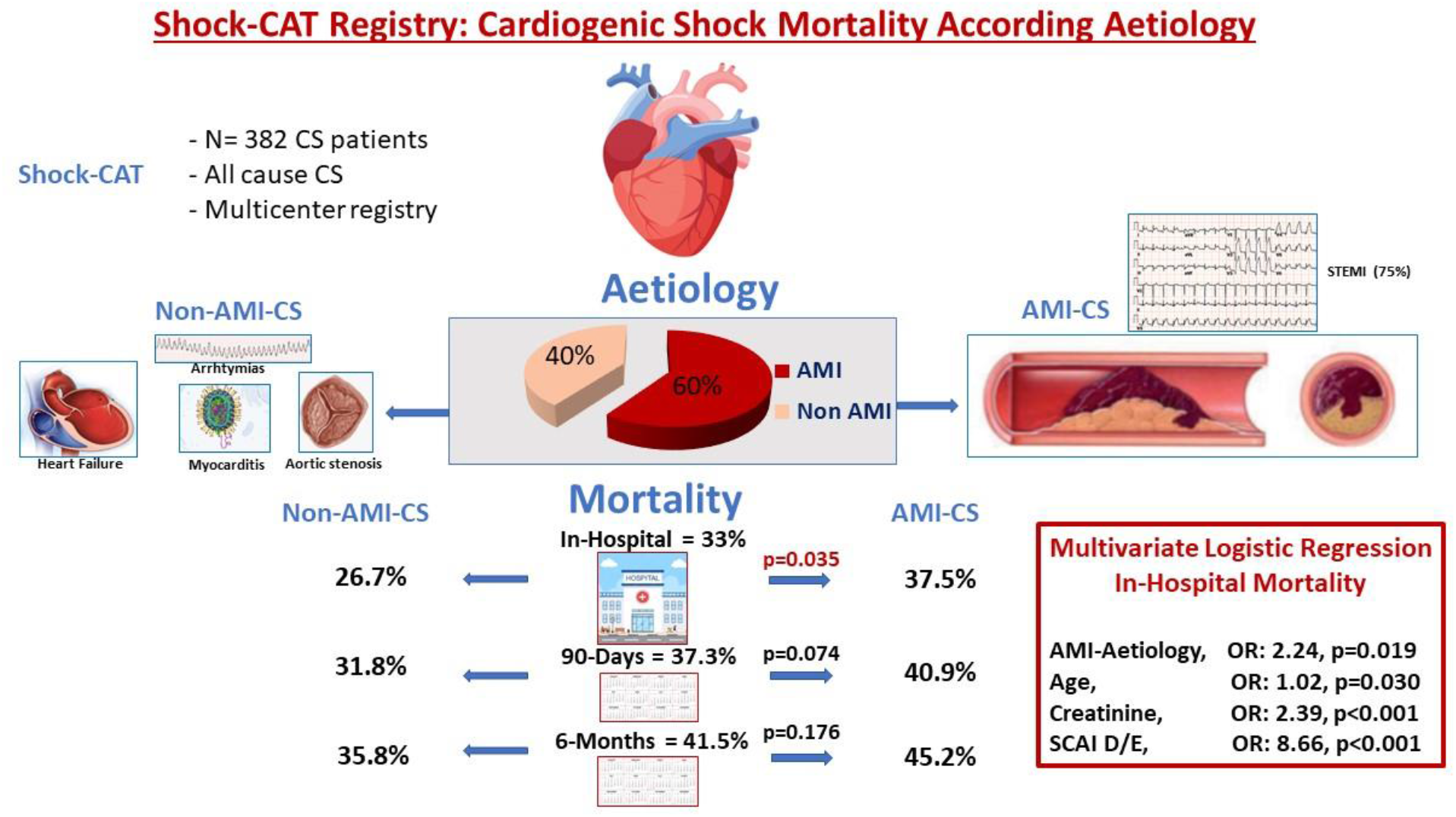
Central Figure. The Shock-CAT registry, main results and summary.

Although there has been a noticeable rise in conditions such as acute heart failure, severe valvulopathies, and cardiac arrest contributing to CS cases within critical care units, AMI remains the predominant cause of CS according to various multicentre registries. This is evident in studies like the CardShock study, where AMI patients constituted 81% of cases (12). Similarly, the recent ECMO-CS randomized trial encompassed 62% AMI cases (18), a proportion consistent with our own registry findings. In our study, discernible distinctions emerge in the clinical characteristics of these two groups. CS patients with ACS tend to be older and possess a higher burden of cardiovascular risk factors. Conversely, among non-AMI CS patients, there is a two-fold increase in the proportion of women, with over 40% having a history of prior heart failure. These observations parallel the outcomes observed in the CardShock trial (12).

Despite overall in-hospital mortality still remains above 33%, this could be representative of real- life CS lethality in the primary PCI era, with extended use of evidence based recommended therapies (1,19,20). All hospitals participating in the Shock-CAT registry were tertiary centers, with specialized ICCU looked after by cardiologist, belonging to a public health system, with a reperfusion network based on primary percutaneous coronary intervention for STEMI patients. Shock-CAT mortality rates was lower than those observed in the IABP-SHOCK II Trial (30-days mortality above 40%) and in the CardShock study (in-hospital mortality 37%) (12), although both studies included patients with similar clinical profile (age, LVEF, aetiology…). Some recent CS studies showed higher mortality than our data, such as the French registry of Puymirat (21) (in- intensive care unit mortality nearby 45% in last period, years 2009-2012). However they included patients with different profile, with only a 12% of ACS patients and 26% of patients being admitted to a non-university hospital. The admission in a tertiary hospital with specialized ICCU has been shown as an independent predictor of lower in-hospital mortality in a recent STEMI CS Spanish registry (22) and the allocation of patients to such centers has been widely recommended (23). In-hospital mortality in the ECMO-CS study was 50% (18), also higher than in our registry, although this trial only included ECMO candidates with refractory CS, limiting direct comparison with other CS registries.

In our registry in-hospital mortality remains higher in AMI-CS patients when compared to non-AMI ones after multivariate adjustment. Consistent with prior reports, our study confirms CS to be a heterogeneous syndrome that includes a broad spectrum of clinical presentation and metabolic disturbances that could imply differences in prognosis, probably due to different phenotypes of CS, as has been previous reported (24,25). Despite the fact that we could not find differences in pH at admission or lactate peak, AMI patients were more often managed with inotropes or MCS than non- ACS patients, even though they had slightly better LVEF than non-AMI-CS. Similar results were observed by Sinha et al. in a recent American single-center CS registry (24).

Non-AMI-CS group included more patients with previous heart failure or myocardial infarction, these factors could suggest a better previous organic accommodation to low cardiac output states, with less pronounced end-organ dysfunction, despite the CS situation. The acute onset of ACS with dramatic decrease of cardiac output, avoids the heart and end-organ preconditioning to this severe hemodynamic scenario and could confer higher mortality to AMI-CS patients. The absence of this preconditioning situation could explain the higher requirement of MCS in AMI-CS patients compared to non-AMI. Similar data were observed in the American study (24), but also in a multicenter European CS registry (26). The use of MCS for treatment of CS patients has been increasing around the world, mainly due to the widespread use of ECMO, although the last results in recent randomized clinical trials (27,28) could influence in future indications. ECMO was implanted in less than 10% of all CS patients, according to 8% reported by Rivas et al in the European registry, but far from 21.9% of the quaternary center of the American’s. These differences are pottentially explained by the nature of the participant centers in our study, all of which are tertiary centers, providing care to all comers, and therefore with a significant elderly or comorbid population being treated but not being candidates for advanced therapies. Moreover, regardless of the lack of evidence-based benefits of IABP when applied routinely in CS patients, it was implanted in nearby 40% of AMI-CS patients in our registry, compared to 44% of the American study or 56% of the European registry. These data confirm that many physicians still believe in the clinical usefulness of IABP in selected patients in the setting of AMI complicated with CS. Similar reasons could explain the use of pulmonary artery catheter in nearby 30% of our patients, meanwhile its use did not impact on mortality in previous study, might help for guiding the management in complex patients (29). The recommendation for the use of MCS only in the setting of “refractory shock” is based on limited available evidence and the appropriate patient selection seems to be important to improve survival overcoming the device-related complications (20).

However, an early and accurate risk stratification seems to be essential for the management of CS, tailoring the invasive therapies such as MCS for high risk patients. CardShock and IABP-SHOCK II scores (12,13) have been widely used and were developed including clinical and classical biochemical parameters (lactate, glucose, renal function…). In the recent years, SCAI classification has been proposed as clinical classification of CS patients. SCAI scale includes 5 stages labelled A-E depending on clinical situation, from A “At risk” of CS to E “Extremis”, that is circulatory collapse (17). SCAI classification has been recently validated is retrospective studies (30) even though further validation in a prospective clinical trial is warranted. The results of our analyses showed IABP-SHOCK II score had better prognostic accuracy than CardShock score for predicting 90-days mortality in our unselected Mediterranean cohort of CS. This result is mainly observed in AMI-CS patients, meanwhile both scores were similar in non-ACS with less discrimination precision. This fact could be explained because IABP was developed in a specific AMI-CS cohort, whereas CardShock included different CS aetiologies. In the European non-selected CS cohort, both scores showed similar discrimination capability and also better results in ACS patients. However, CardShock and IABP scores do not have very high predictive accuracy, with an area under the curve nearby 0.75 in the original CS cohorts. Similar AUC results have been reproduced in our registry and although IABP-SHOCK II had better discrimination than CardShock, the whole values around 0.62-0.74 seem to be suboptimal for an accurate risk stratification. That’s why it could be mandatory to find alternative risk stratification strategies, such as biomarkers or protein-based risk scales. Rueda et al. observed that the combination of 4 systemic circulating proteins (CS4P model) improved the overall performance predictive metrics of CardShock and IABP-SHOCK II score, with a net reclassification improvement (31). This could be a promising future approach to CS risk stratification.

## Study limitations

The main limitation of this study could be the limited size of the cohort included, this fact could restrict the conclusions. However, the multicenter inclusion with similar center capabilities confers great value to these real-life results, which could be representative of CS prognosis. CS management was performed following the local physiciańs criteria, although all patients were admitted in a specialized ICCU with standardized evidence-based therapies, e.g. reperfusion with primary PCI was mandatory for STEMI patients, following the reperfusion network rules of a public health system. Shock-CAT registry was conducted in several ICCU prioritizing cardiac scores for risk prediction, while information about general intensive care severity risk scores such as APACHE or SOFA scores were not available in our registry. Finally, IABP-SHOCK II risk score marks depending on the final TIMI grade after revascularization, although in non-AMI CS group there were 31% of patients without coronary angiography.

## CONCLUSIONS

In an unselected cohort of CS cases from the Mediterranean region, AMI continues to account for over 60% of cases. Approximately one-third of CS patients experienced in-hospital mortality, with AMI-CS patients facing twice the in-hospital mortality rate compared to non-AMI patients. The trend of all-cause mortality remains elevated in AMI cases compared to non-AMI cases at both the 90-day and 6-month marks. When assessing the accuracy of predicting 90-day mortality, the IABP-SHOCK II score exhibit greater precision for AMI patients than for those with non-AMI-CS, while the CardShock score demonstrates a similar level of accuracy in both groups.

## Data Availability

The data are available if someone needs it. All data are under the own of the correspondence author

## Acknowledgements

The authors thank all members of the Intensive Cardiac Care Units working group of Catalan Society of Cardiology to their contribution in this registry.

---

**Supplementary Table 1.** ICCU medical therapies.

|  | All patients<br>(n=382) | AMI-CS<br>(n=232) | Non-AMI-CS<br>(n=150) | P Value |
| --- | --- | --- | --- | --- |
| **Pharmacological treatment in ACCU** |  |  |  |  |
| Dobutamine, n (%) | 297 (79.2) | 197 (86.4) | 100 (68) | <0.001 |
| Norepinephrine, n (%) | 291 (77.6) | 181 (79.4) | 110 (74.8) | 0.302 |
| Epinephrine, n (%) | 74 (19.7) | 51 (22.4) | 25 (15.6) | 0.110 |
| Dopamine, n (%) | 6 (1.6) | 5 (2.2) | 1 (0.7) | 0.254 |
| Nitrates, n (%) | 131 (35.9) | 83 (37.1) | 48 (34.0) | 0.779 |
| Nitroprusside, n (%) | 88 (23.8) | 48 (21.2) | 40 (27.8) | 0.150 |
| Levosimendan, n (%) | 22 (8.5) | 13 (5.7) | 19 (12.9) | 0.015 |
| Diuretics, n (%) | 330 (88.7) | 194 (85.8) | 136 (93.2) | 0.030 |
| Aspirin, n (%) | 306 (82) | 232 (100) | 78 (53.8) | <0.001 |
| Clopidogrel, n (%) | 177 (47.7) | 146 (64.6) | 31 (21.4) | <0.001 |
| Ticagrelor, n (%) | 75 (20.3) | 71 (31.4) | 4 (2.8) | <0.001 |
| Prasugrel, n (%) | 21 (5.7) | 20 (8.9) | 1 (0.7) | 0.001 |
| Glycoprotein IIb/IIIa inhibitors, n (%) | 35 (9.9) | 132 (14.9) | 3 (2.2) | <0.001 |
| Any heparin, n (%) | 319 (86.4) | 195 (86.3) | 124 (86.7) | 0.906 |
| Low-molecular-weight heparin, n (%) | 187 (50.7) | 106 (46.9) | 81 (56.6) | 0.068 |
| Unfractionated heparin, n (%) | 173 (46.9) | 120 (53.1) | 53 (37.1) | 0.003 |
| Amiodarone, n (%) | 167 (45) | 101 (44.5) | 66 (45.8) | 0.800 |
| Lidocaine, n (%) | 22 (5.9) | 19 (8.4) | 3 (2.1) | 0.012 |
| Procainamide, n (%) | 18 (4.9) | 8 (3.5) | 10 (6.9) | 0.135 |
| Any antiarrhythmic drugs, n (%) | 180 (48.5) | 105 (46.3) | 75 (52.1) | 0.274 |
| Digoxin, n (%) | 41 (11.1) | 19 (8.4) | 22 (15.3) | 0.039 |
| ACEI/ARB, n (%) | 204 (54.5) | 122 (53.5) | 82 (56.2) | 0.615 |
| Espironolactone/eplerenone, n (%) | 146 (39) | 89 (39) | 57 (39) | 0.990 |
| β-Blockers, n (%) | 196 (52.5) | 114 (50.2) | 82 (56.2) | 0.262 |
| Ivabradine, n (%) | 39 (10.6) | 25 (11.2) | 14 (9.7) | 0.662 |
| Calcium channel blockers, n (%) | 36 (9.7) | 20 (8.8) | 16 (11.0) | 0.471 |
| Statins, n (%) | 286 (76.9) | 199 (87.3) | 87 (60.4) | <0.001 |

ICCU: Intensive Cardiac Cares Unit; ACEI: Angiotensin Convertase Enzyme Inhibitor; ARB: angiotensin
Receptor Blocker

**Supplementary Table 2:** Baseline characteristics for sensitivity analysis after propensity Score matching.

|  | All patients<br>(n=208) | AMI-CS<br>(n=104) | Non-AMI-CS<br>(n=104) | P Value |
| --- | --- | --- | --- | --- |
| **Demographics** |  |  |  |  |
| Age, mean (SD) | 64.0 (14.7) | 63.3 (13.1) | 64.7 (16.2) | 0.500 |
| Gender, female, n (%) | 74 (35.6) | 34 (32.7) | 40 (38.5) | 0.385 |
| BMI, kg/m<sup>2</sup> mean (SD) | 27.2 (4.7) | 27.6 (4.3) | 26.9 (5.1) | 0.299 |
| **History, n (%)** |  |  |  |  |
| Smoking | 58 (27.9) | 36 (34.6) | 22 (21.2) | 0.060 |
| Dyslipidaemia | 125 (60.1) | 63 (60.9) | 62 (59.6) | 0.347 |
| Hypertension | 131 (63.0) | 68 (65.4) | 63 (60.6) | 0.241 |
| Diabetes mellitus | 76 (36.5) | 37 (35.6) | 39 (37.5) | 0.358 |
| Liver disease | 9 (4.3) | 5 (4.8) | 4 (3.8) | 0.733 |
| Cerebrovascular disease | 17 (8.3) | 7 (6.8) | 10 (9.7) | 0.447 |
| Peripheral arterial disease | 25 (12.1) | 12 (11.7) | 13 (12.6) | 0.831 |
| Chronic kidney disease | 27 (13.0) | 17 (16.7) | 10 (9.6) | 0.352 |
| COPD | 26 (12.7) | 13 (12.9) | 13 (12.5) | 0.936 |
| Neoplasia | 23 (11.2) | 9 (9.7) | 14 (13.6) | 0.269 |
| Anaemia | 44 (21.2) | 21 (20.2) | 23 (22.1) | 0.734 |
| Previous heart failure | 27 (13.0) | 9 (8.7) | 18 (17.3) | 0.063 |
| Previous MI | 37 (17.8) | 19 (18.3) | 18 (17.3) | 0.856 |
| Previous PCI | 25(12.0) | 13 (12.5) | 12 (11.5) | 0.977 |
| Previous CABG | 9 (4.3) | 4 (3.8) | 5 (4.8) | 0.944 |
| Previous valvular surgery | 11 (5.3) | 1 (1.0) | 10 (9.6) | 0.013 |
| Pacemaker carrier | 10 (4.8) | 2 (1.9) | 8 (7.7) | 0.092 |
| ICD carrier | 5 (2.4) | 0 (0) | 5 (4.8) | 0.046 |
| Previous ablation | 4 (1.9) | 0 (0) | 4 (3.8) | 0.076 |
| **Clinical presentation, mean (SD)** |  |  |  |  |
| Systolic blood pressure, mmHg | 93.5 (28.1) | 91.5 (28.4) | 95.5 (27.9) | 0.311 |
| Diastolic blood pressure, mmHg | 58.3 (18.7) | 58.2 (18.9) | 58.5 (18.5) | 0.913 |
| Heart rate, bpm | 92.9 (31.3) | 92.9 (32) | 92.9 (30) | 0.995 |
| LVEF, % | 31.9 (13.9) | 32.3 (13.1) | 31.5 (14.7) | 0.694 |
| Haemoglobin, g/dL | 12.7 (2.2) | 12.7 (2.5) | 12.8 (2.0) | 0.761 |
| Glucose, mg/dL | 212 (137) | 214 (110) | 209 (160) | 0.795 |
| Creatinine, mg/dL | 1.56 (0.9) | 1.54 (1.0) | 1.57 (0.8) | 0.835 |
| eGFR<sub>CKD-EPI</sub>, mL/min/1.73 m<sup>2</sup> | 58.1 (30.6) | 61.2 (33.2) | 55.1 (27.6) | 0.157 |
| pH, at admission | 7.27 (0.17) | 7.26 (0.16) | 7.28 (0.18) | 0.543 |
| Lactate peak, mmol/L | 5.9 (5.5) | 5.3 (3.9) | 6.6 (6.7) | 0.090 |
| Shock index | 1.2 (0.6) | 1.1 (0.6) | 1.2 (0.6) | 0.305 |
| **ECMO Capability hospital, n (%)** | **156 (75)** | **77 (74)** | **79 (76)** | **0.749** |

STEMI, ST-segment elevation myocardial infarction; NSTEMI, non-ST-segment elevation myocardial infarction; BMI, body mass index; MI, myocardial infarction; PCI, percutaneous coronary intervention; CABG, coronary artery bypass graft; LVEF, left ventricular ejection fraction; eGFR<sub>CKD-EPI</sub>, estimated glomerular filtration rate by the Chronic Kidney Disease Epidemiology Collaboration formula. COPD: chronic obstructive pulmonary disease; ECMO: Extracorporeal membrane oxygenation; ICD: Implantable cardiac defibrillator; SD: standard deviation.

